# Compound climate extremes predict dengue risk at operational lead times across Brazil

**DOI:** 10.64898/2026.10.16.26365716

**Authors:** Chloe Fletcher, Carles Milà, Giovenale Moirano, Eduardo Araujo, Daniela Lührsen, Claudia T. Codeço, Flávio Codeço Coelho, Rachel Lowe

**Affiliations:** Barcelona Supercomputing Center (BSC), Barcelona, Spain; Department of Medicine & Life Sciences, Universitat Pompeu Fabra, Barcelona, Spain; Department of Medical Sciences, University of Turin, Turin, Italy; School of Applied Mathematics, Fundação Getulio Vargas, Rio de Janeiro, Brazil; Programa de Computação Científica, Fundação Oswaldo Cruz, Rio de Janeiro, Brazil; Catalan Institution for Research and Advanced Studies (ICREA), Barcelona, Spain; London School of Hygiene and Tropical Medicine, London, UK; Saw Swee Hock School of Public Health, National University of Singapore, Singapore

## Abstract

Brazil has one of the highest dengue burdens globally, with transmission influenced by climate events. Yet, the effects of compound climate extremes on dengue risk and prediction have not been quantified to date. Using a Bayesian spatiotemporal model of monthly dengue cases across 435 health regions from 2010 to 2025, we estimated relative risk associated with interacting long-lag and short-lag drought indicators and temperature anomalies across nine Köppen-Geiger climatic zones, adjusting for absolute temperature and El Niño-Southern Oscillation. Long-lag dry, mid-lag warm and short-lag wet conditions were associated with the highest relative risk in five climatic zones, with temperature anomalies modifying hydrometeorological dynamics more strongly in temperate than tropical areas. Climate covariates improved the prediction of case counts and outbreaks compared to baseline models at lead times of 1, 3, 6 and 12 months, and for the upcoming season 5 months ahead. Improvements were greatest in the Northeast, Southeast, and Midwest. Recent dengue incidence provided the greatest improvement at the 1-month lead, whereas combining incidence with climate information performed best at 3- and 6-month leads. We extended our framework to weekly state-level predictions of the season ahead to align with national public health planning. We proposed a multi-staged dengue forecasting approach to enhance early warning and response across Brazil.

## Introduction

Dengue fever is a febrile illness caused by infection with the dengue virus, transmitted primarily by *Aedes aegypti* mosquitoes, with *Aedes albopictus* serving as a secondary vector (Lambrechts et al., 2010). Around half of the global population are currently at risk of dengue (World Health Organization, 2025). Projections indicate that climate change will place an additional 3.6 billion people at risk by 2067 relative to the 1970-1999 baseline (Colón-González, Sewe, et al., 2021). In 2024, there was an unprecedented surge in dengue infections with 14.1 million cases recorded worldwide (Haider et al., 2025). This represented a 12-fold increase compared to 2014 and more than double the previous record of 6.4 million cases in 2023. The Americas reported the greatest burden, accounting for 92% of global cases, of which Brazil reported 10.3 million infections (Haider et al., 2025). This rapid increase coincided with the 2023-2024 El Niño event, which exacerbated extreme heat, rainfall and drought across the region (Arguez et al., 2025).

Climatic conditions influence dengue transmission both directly and indirectly. Temperature affects mosquito development, survival and biting rates, as well as the extrinsic incubation period (Mordecai et al., 2019). Transmission of dengue virus by *Aedes aegypti* is predicted to occur between 17.8°C (95% credible interval, CrI, 14.6-21.2) and 34.5°C (95% CrI 34.1-35.8), with an optimum at 29.1°C (95% CrI 28.5-29.8) (Mordecai et al., 2019). Rising average temperatures are expanding the geographic suitability of *Aedes* mosquitoes into higher latitudes and altitudes (Ryan et al., 2019). However, the effects of hydrometeorological events on dengue can be more complex. Heavy rainfall can increase standing water that create or sustain mosquito breeding sites, or flush larvae from containers (Koenraadt & Harrington, 2008; Stewart Ibarra et al., 2013). Droughts promote household water storage that serve as ideal habitats if poorly maintained (Gómez-Vargas et al., 2024; Lowe et al., 2018; Stewart Ibarra et al., 2013). When extreme weather events occur in succession, these hazards can compound, creating cascading effects on the economy, healthcare systems, infrastructure, and human behaviour that can amplify exposure to vectors and erode resilience (Semenza et al., 2022).

Brazil spans diverse contexts, from small communities in the Amazon rainforest to densely-populated megacities, across tropical, semi-arid and temperate climates (Peel et al., 2007). This heterogeneity can foster complex transmission dynamics (Gouveia et al., 2025). Historically, the South and Amazon regions experienced limited spread of dengue, but increased warming, urbanisation, and connectivity between endemic areas have eroded these barriers and driven expansion into new territories (Barcellos et al., 2024; C. T. Codeco et al., 2022; Lee et al., 2021). Hydrometeorological extremes have demonstrated delayed and nonlinear effects on dengue incidence, with increased relative risk 0-3 months after extremely wet conditions or 3-5 months after extremely dry conditions (Lowe et al., 2021). When interacted with urbanisation level, extreme rainfall represented a greater risk in less urbanised areas, whereas drought showed the greatest risk in highly urbanised areas. Similar findings on the delayed effects of climate extremes on dengue have been found in Barbados (Lowe et al., 2018), Peru (Mills & Donnelly, 2024), and Vietnam (Gibb et al., 2023).

Many climate-related impacts arise from combined rather than individual events (Zscheischler et al., 2018). Compound events describe multiple climatic hazards that jointly contribute to societal risk, classed into four (non-mutually-exclusive) types: (1) preconditioned, (2) multivariate, (3) temporally compounding, and (4) spatially compounding (Zscheischler et al., 2020). Concurrent events may be classed as multivariate (i.e. joint impact in the same location) or spatially compounding (i.e. joint impact across connected locations), whereas sequential events may be classed as preconditioned (i.e. a prior condition or event amplifies the impact of a subsequent event) or temporally compounding (i.e. joint impact from successive events in the same location), depending on the mechanism. For dengue, concurrent heat and rainfall extremes were associated with increased relative risk compared to moderate or isolated events across four countries in South and Southeast Asia (Y. Wang et al., 2024). By contrast, concurrent drought and heatwave events decreased dengue risk compared to either hazard individually over seven Southeast Asian countries (D. Wang et al., 2026). In Vietnam, dengue risk was highest following sequential dry-then-wet conditions in seven of eight subregions, where the magnitude varied across emerging and endemic transmission settings (Fletcher et al., 2026). In Barbados, three sequential weather events were found to compound dengue risk, with the highest incidence associated with dry-then-warm-then-wet conditions (Fletcher et al., 2025). The effects of compound climate extremes on dengue risk have not yet been quantified in Brazil, with limited studies across diverse spatial contexts or tested within a predictive framework.

Anticipating the timing, location and likelihood of outbreaks in advance can help public health decision-makers determine when, where and how to mobilise resources most effectively, reducing both health and economic impacts. The financial costs of *Aedes*-borne diseases from 1975-2020 was estimated at US$94.7 billion globally, where Brazil represented the second-highest contributor, spending $12.0 billion on dengue alone (Roiz et al., 2024). Loss and damage expenditure was estimated to be 11-times higher than for disease management, highlighting the need to move from reactive to proactive response. Predictive models offer advanced warning of changing dengue risk patterns, with climate forecast integration extending lead times by several months. For the World Cup 2014 in Brazil, a Bayesian spatiotemporal model combined with seasonal climate forecasts produced probabilistic predictions of dengue risk per microregion three months ahead (Lowe et al., 2014), outperforming a null model based on historical averages (Lowe et al., 2016). Since 2015, the Infodengue early warning system has been operational, combining multi-sourced epidemiological, climatic and demographic data to generate municipality-level risk indicators across Brazil (C. Codeco et al., 2018). The system conducts nowcasting by correcting for reporting delays (Bastos et al., 2019), and detects sustained transmission by estimating the dengue reproduction number (Codeço et al., 2018). In recent years, the Infodengue and Mosqlimate teams have launched annual forecasting challenges, where international teams generate weekly state-level dengue forecasts for a whole season several weeks in advance, to form a weighted ensemble model (Araujo et al., 2026). These predictions are shared with the Brazilian Ministry of Health to aid public health preparedness for the upcoming season.

Despite these advances, challenges persist in generating forecasts that are both reliable and useful in operational contexts. The value added by climate information depends on the lead time, with recent surveillance enhancing skill at shorter lead times and climate covariates at longer leads (Beal et al., 2025). Weekly predictions align best with surveillance and decision-making, yet climate forecasts are more skilful at monthly to seasonal timescales (White et al., 2017). Whilst longer lead times provide more opportunity to carry out preventative public health interventions, seasonal forecast skill declines as this time horizon increases. This skill can also be spatially heterogeneous due to differences in transmission regimes, surveillance practices, and climate-driven dengue dynamics (Almeida et al., 2022; Araujo et al., 2026; Fletcher et al., 2026). Improved understanding of these complexities can enhance the design of robust climate-informed predictive models that maximise forecast accuracy and practical utility. This study evaluates how predictive skill varies across operational lead times for a dengue risk model in Brazil. We present a Bayesian spatiotemporal modelling framework that captures the effects of compound climate extremes, as well as absolute temperature and El Niño-Southern Oscillation (ENSO), on monthly health region level dengue risk across different climatic zones. We evaluate predictive skill at rolling lead times of 1, 3, 6 and 12 months, and for the upcoming dengue season several months in advance. The model is extended to the weekly resolution to align with public health preparedness and response planning in Brazil.

## Results

### Relationships between compound climate extremes and dengue risk vary across climatic zones

We fit Bayesian spatiotemporal models to monthly dengue case counts across 435 health regions in Brazil from January 2010 to May 2025 (Figure 1), evaluating climate covariates, lags and effect stratifications for improved goodness-of-fit and credible effect sizes. The best-fitting model (M_1_) included a three-way interaction between the 12-month Standardized Precipitation Evapotranspiration Index (SPEI-12; lagged 2 months), 3-month SPEI (SPEI-3; lagged 0 months) and 6-month standardised mean temperature anomaly (lagged 1 month), as well as additive terms for the 6-month absolute mean temperature (lagged 1 month) and the 3-month mean Niño 3.4 sea surface temperature (SST) anomaly (lagged 5 months) (Figure 2). The effects of ambient temperature and drought indicators were stratified by the nine Köppen-Geiger climatic zones of Brazil using random slopes, whereas the SST anomaly was modelled as a smooth nonlinear effect. The model additionally included a monthly cyclic second-order random walk (RW2) random effect replicated by state, a yearly independent and identically distributed (IID) random effect for each dengue season, defined as October to September, and a spatial random effect on the health region neighbourhood matrix using a modified Besag-York Mollié (BYM2) model.

**Figure 1.**
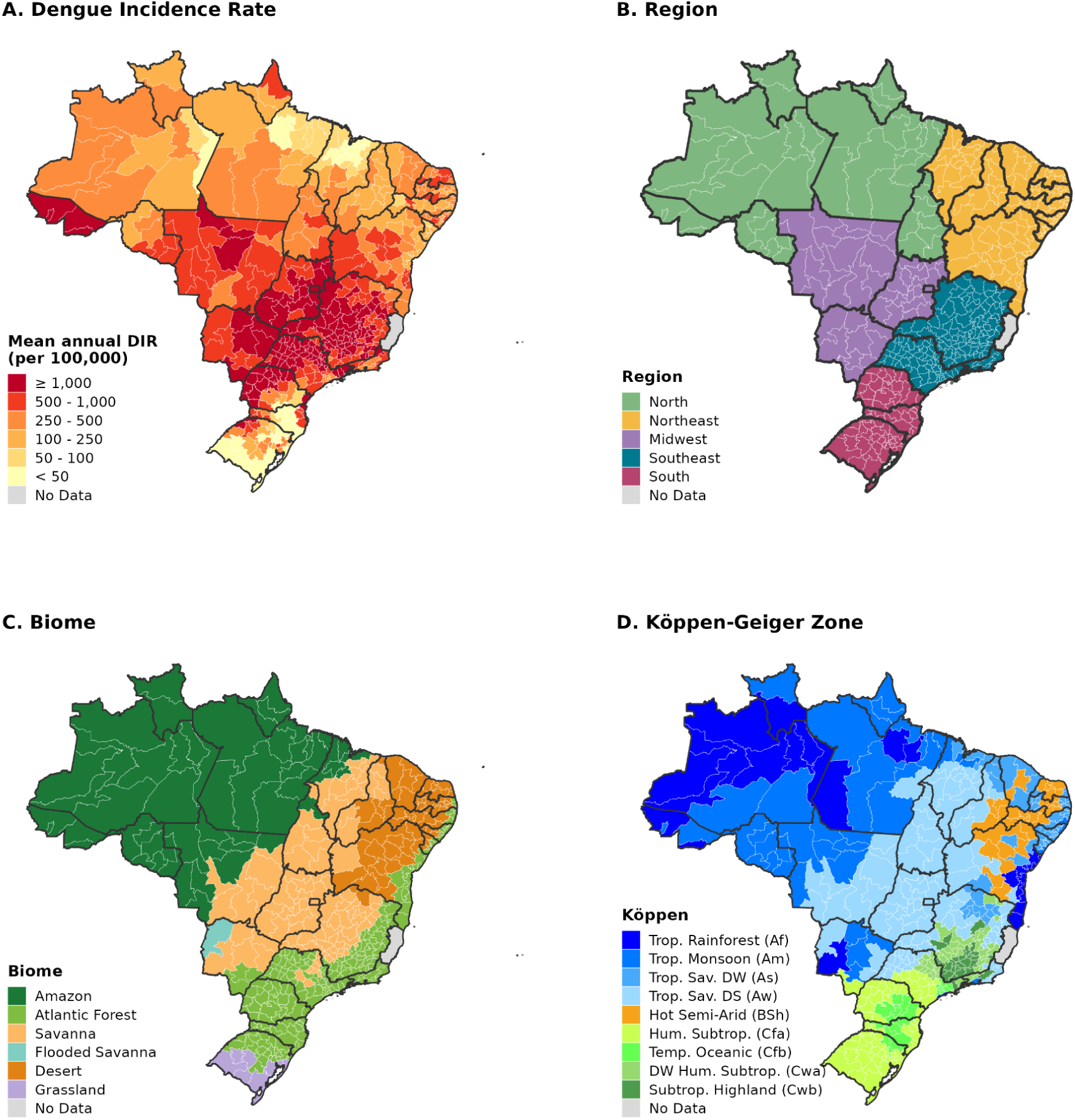
Dengue incidence and spatial classification of Brazilian health regions. (A) Mean annual dengue incidence (DIR) per 100,000 population for 435 health regions from January 2010 to May 2025. (B) Macro-region classification of health regions. (C) Biome classification of health regions, assigned from the dominant municipality-level classification. (D) Köppen-Geiger climate zone classification of health regions into tropical (Af, Am, As, Aw), semi-arid (BSh), and temperate (Cfa, Cfb, Cwa, Cwb) zones, assigned from the dominant municipality-level classification. “Trop.” = Tropical, “Subtrop.” = Subtropical, “Temp” = Temperate, “Sav.” = Savannah, “Hum.” = Humid, “DW” = Dry Winter, “DS” = Dry Summer. Thin lines delineate health regions and thick lines delineate federal units. Health regions in the state of Espírito Santo are shown in grey due to lack of available surveillance data.

**Figure 2.**
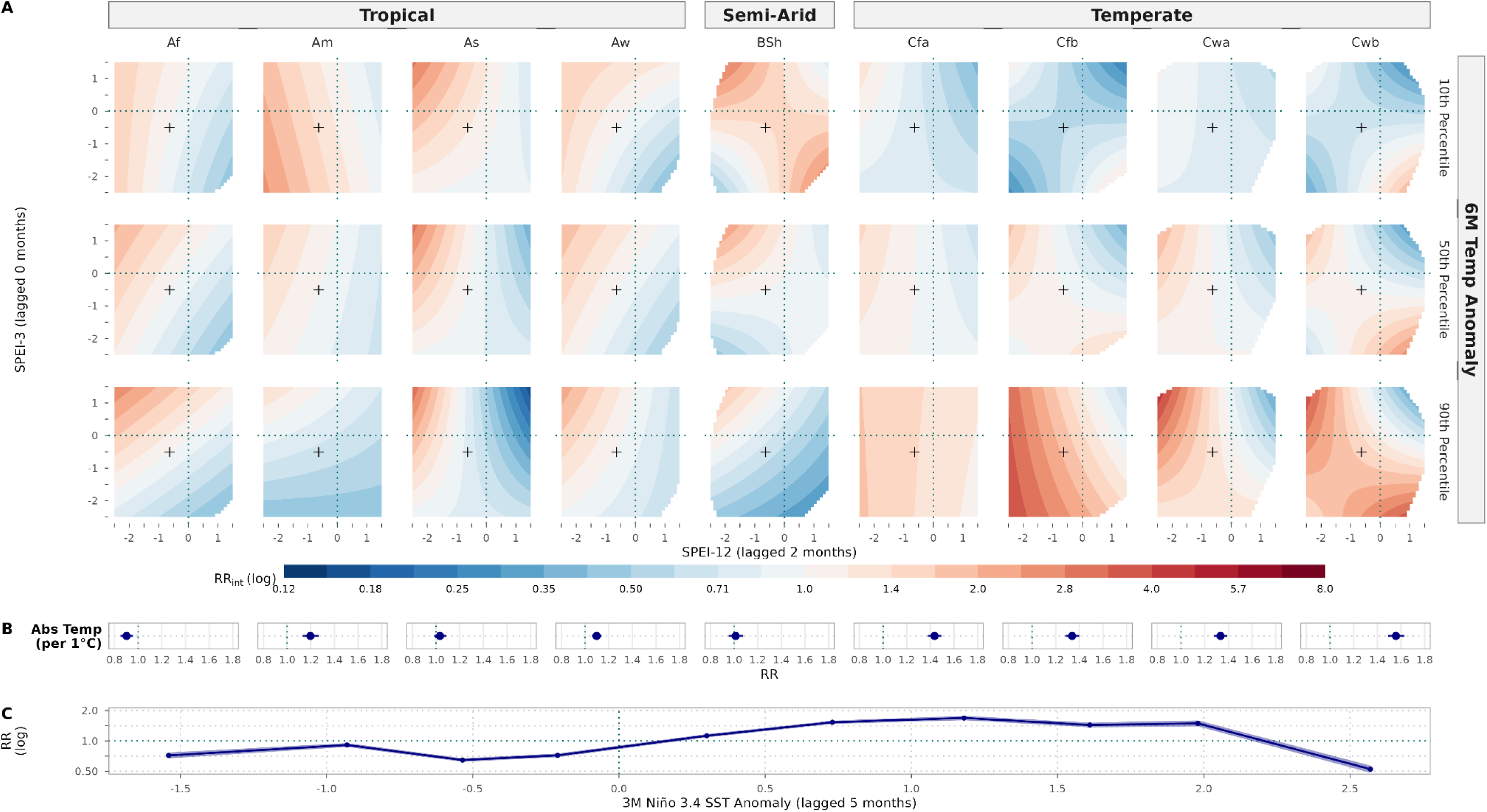
The effect of compound climate extremes, absolute temperature and ENSO on dengue relative risk across climatic zones. Risks are quantified for the best-fitting model (MJ for covariate effects stratified by nine Köppen-Geiger zones (columns) grouped into first-level classifications: tropical, semi-arid, and temperate. (A) Relative risk of dengue (RR_mt_) for the three-way interaction between the SPEI-12 (lagged 2 months, x-axis). SPEI-3 (lagged 0 months, y-axis). and the 6-month standardised mean temperature anomaly (lagged 1 month) shown at the 10th, 50th and 90th percentiles (rows). Risk is expressed relative to the national median conditions over the study period (SPEI-12 = −0.64, SPEI-3 = −0.51, 6M Temp. Anom. = 1.00) marked by a cross, and have exclusively been calculated on the observed range of SPEI-12 and SPEI-3 values. Dotted lines indicate the zero anomaly for each index. Colours follow the log scale. (B) Relative risk per 1°C increase in the 6-month mean temperature (lagged 1 month), showing the mean and 95% credible interval. (C) Nonlinear effect of the 3-month mean Niño 3.4 SST anomaly (lagged 5 months), showing the mean (line) and 95% credible interval (shading)

We calculated the dengue relative risk (RR_int_) associated with compound climate extremes across 9 climatic zones relative to national median conditions over the study period (i.e. SPEI-12 = −0.64, SPEI-3 = −0.51, temperature anomaly = 1.00) (Figure 2A). For each zone, we identified the combination of extremes associated with the maximum and minimum RR_int_, estimated across the typical range of observations (2.5th to 97.5th percentile) for each covariate in that zone (Table 1). The maximum RR_int_ was associated with long-lag dry conditions (i.e., negative SPEI-12) in 8 of 9 climatic zones, compounded by warmer temperature anomalies (i.e. positive 6-month temperature anomalies) in 7 zones, and by short-lag wet conditions (i.e., positive SPEI-3) in 5 zones. The compound sequence of dry followed by warm and wet events (dry-warm-wet) was associated with the maximum RR_int_ in 5 climatic zones (Table 1), but increased RR_int_ in all zones (Figure 2A). In temperate areas, warmer conditions were associated with increased dengue RR_int_, compounding dry-then-wet conditions to give the maximum RR_int_ in temperate zones with a dry winter (Cwa, Cwb), and by dry-then-dry (i.e. persistently dry) conditions in temperate zones without a dry season (Cfa, Cfb). Semi-arid areas (BSh) showed the maximum RR_int_ with compound wet-cool-dry conditions. Most tropical climates (Af, Aw, As) showed the maximum RR_int_ with dry-warm-wet conditions, except tropical monsoon (Am) areas. However, long-lag and short-lag SPEI interactions in all tropical zones exhibited consistent dengue risk patterns across temperature anomalies, indicating a less pronounced modification effect.

**Table 1.** Minimum and maximum dengue relative risk associated with compound climate extremes across climatic zones. Minimum and maximum relative risk (RR_int_) of dengue, mean and 95% credible interval, for the three-way interaction between the 6-month standardised mean temperature anomaly (lagged 1 month), SPEI-12 (lagged 2 months), SPEI-3 (lagged 0 months) stratified by nine Köppen-Geiger climatic zones, estimated for the best-fitting model (M_1_). Within each zone, the RR_int_ was calculated between the 2.5th and 97.5th percentiles of observations for each covariate. Risks are quantified relative to national median conditions over the study period (SPEI-12 = −0.6, SPEI-3 = −0.5, Temp. Anom. = 1.0).

| Köppen | Dengue Risk | $RR_{int}$ [95% CrI] | Temp. Anom. | Long SPEI-12 | Short SPEI-3 |
| --- | --- | --- | --- | --- | --- |
| Af | Minimum | 0.40 [0.27, 0.68] | 4.39 | 1.32 | -2.21 |
| Af | Maximum | 3.77 [2.40, 6.14] | 4.39 | -2.15 | 1.72 |
| Am | Minimum | 0.23 [0.19, 0.28] | 4.04 | -2.49 | -2.20 |
| Am | Maximum | 2.88 [2.10, 3.81] | -0.66 | -2.49 | -2.20 |
| As | Minimum | 0.13 [0.10, 0.19] | 2.65 | 1.54 | 1.89 |
| As | Maximum | 3.01 [2.38, 3.87] | 2.65 | -2.22 | 1.89 |
| Aw | Minimum | 0.47 [0.40, 0.54] | -0.55 | 0.73 | -2.28 |
| Aw | Maximum | 2.45 [1.96, 3.09] | 4.15 | -2.68 | 1.05 |
| BSh | Minimum | 0.37 [0.27, 0.55] | 2.45 | 1.23 | -2.14 |
| BSh | Maximum | 4.87 [3.49, 6.60] | -1.26 | 1.23 | -2.14 |
| Cfa | Minimum | 0.24 [0.19, 0.31] | -1.25 | 1.69 | 1.91 |
| Cfa | Maximum | 2.31 [2.00, 2.69] | 3.83 | -2.51 | -2.26 |
| Cfb | Minimum | 0.16 [0.10, 0.21] | -1.26 | 1.60 | 1.76 |
| Cfb | Maximum | 6.35 [4.90, 8.52] | 3.45 | -2.23 | -2.12 |
| Cwa | Minimum | 0.59 [0.38, 0.99] | 4.14 | 0.62 | 1.11 |
| Cwa | Maximum | 7.99 [5.71, 10.59] | 4.14 | -2.43 | 1.11 |
| Cwb | Minimum | 0.35 [0.26, 0.42] | -0.81 | 0.67 | 1.10 |
| Cwb | Maximum | 4.55 [3.10, 5.98] | 3.79 | -2.17 | 1.10 |

The 6-month absolute mean temperature was associated with increased dengue relative risk (RR) in 8 of 9 climatic zones (Figure 2B). Each 1°C warming was associated with the highest RR across temperate zones (RR = 1.56, 95% CrI 1.49-1.63 in Cwb; RR = 1.44, 95% CrI 1.38-1.49 in Cfa), and with smaller increases across tropical climates (Aw, Am). Higher temperatures in tropical rainforest (Af) areas were associated with decreased dengue risk (RR = 0.90, 95% CrI 0.85-0.96), with null effects elsewhere. Dengue risk was also associated with the ENSO phase captured by the nonlinear SST anomaly (Figure 2C), decreasing relative to neutral conditions across all negative anomalies, and increasing from 0.3-2.2°C (maximum at 1.2°C with RR = 1.69, 95% CrI 1.61-1.78), above which dengue risk decreased.

### Climate information improves predictive skill across all operational lead times

We simulated out-of-sample prediction for lead times of 1, 3, 6 and 12 months using rolling-origin cross-validation with an expanding window from October 2020 to May 2025. The best-fitting model (M_1_) was evaluated alongside three other formulations: the same model excluding the yearly random effect (M_2_), and two corresponding random-effects-only baseline models (B_1_ and B_2_). Predictive skill was evaluated across all months and health regions using three metrics. The Continuous Ranked Probability Score (CRPS) measures how closely the posterior predictive distribution matches the observed cases counts (case estimation). The Brier Score (BS) measures the accuracy of probabilistic predictions (probabilistic accuracy). The Area Under the Receiver Operating Characteristic Curve (AUC) measures how well the predicted outbreak probabilities discriminate between outbreaks and non-outbreak months (outbreak detection). Lower CRPS and BS, and higher AUC, indicate better predictive skill.

The best-fitting model (M_1_) demonstrated the strongest overall predictive skill (Figure 3; Table 2). For case estimation, M_1_ performed best at lead times of 1 and 3 months, whereas its derivative excluding the yearly random effect (M_2_) performed best at 6- and 12-month leads according to the CRPS. M_1_ outperformed all models at all lead times for probabilistic accuracy and outbreak detection, with the skill of M_2_ converging at the 12-month lead for both the Brier Score and AUC. Similar results were found when comparing baseline models (B_1_ vs B_2_). These findings demonstrate that inclusion of the yearly random effect degraded skill more rapidly as lead time increased, although skill decreased with lead time for all models. For M_1_, the CRPS decreased by 6.9%, BS by 18.7% and AUC by 0.08 from lead 1 to 12. For M_2_, decreases were by 4.4%, 11.5% and 0.05 respectively.

**Figure 3.**
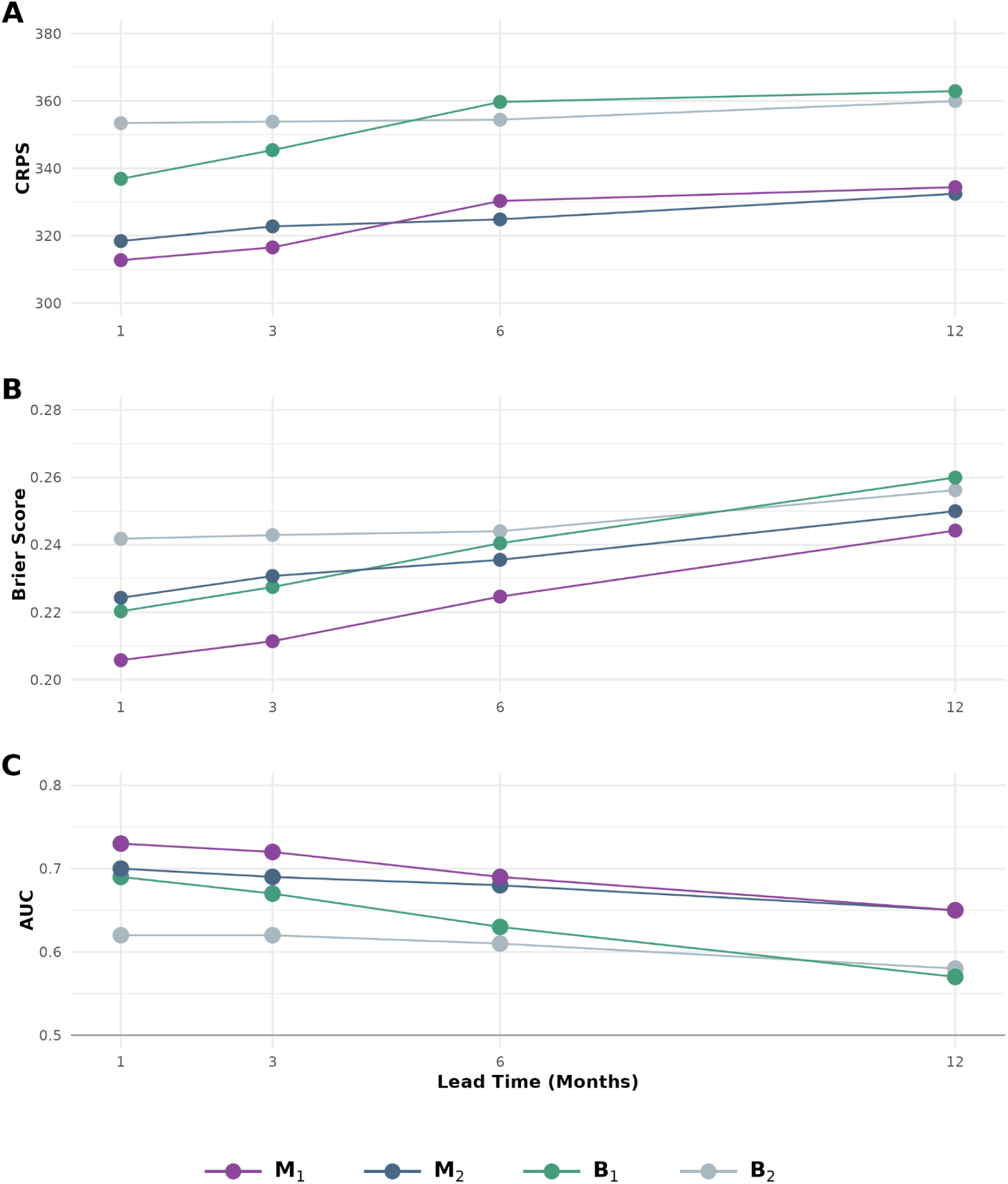
Predictive skill for case estimation, probabilistic accuracy and outbreak detection across operational lead times. Out-of-sample predictive skill at lead times of 1, 3, 6 and 12 months, evaluated across all months and health regions using a rolling-origin cross-validation with an expanding window from October 2020 to May 2025. Four model formulations were compared: the best-fitting climate-informed model (M_1_), the same without the yearly random effect (M_2_), and the corresponding random-effects-only baselines (B_1_ and B_2_). (A) Continuous Ranked Probability Score (CRPS) for case estimation, (B) Brier Score for probabilistic accuracy, (C) Area Under the Receiver Operating Characteristic Curve (AUC) for outbreak detection. Lower CRPS and Brier Score, and higher AUC, indicate better skill. Outbreaks were defined as dengue incidence above the historical 75th percentile for the calendar month and health region.

**Table 2.** Predictive skill for case estimation, probabilistic accuracy and outbreak detection across operational lead times. Out-of-sample predictive skill at lead times of 1, 3, 6 and 12 months, evaluated across all months and health regions using rolling-origin cross-validation with an expanding window from October 2020 to May 2025.Four model formulations were compared: the best-fitting climate-informed model (M_1_), the same without the yearly random effect (M_2_), and the corresponding random-effects-only baselines (B_1_ and B_2_). The Continuous Ranked Probability Score (CRPS) measures case estimation, the Brier Score measures probabilistic accuracy, the Area Under the Receiver Operating Characteristic Curve (AUC) measures outbreak detection. Lower CRPS and Brier Score, and higher AUC, indicate better skill. Trigger denotes the minimum outbreak probability to raise an alert, derived from the ROC curve as the top-left value. The True Positive Rate (TPR) and False Positive (FPR) are estimated for Trigger. Outbreaks were defined as dengue incidence above the historical 75th percentile for the calendar month and health region.

| Model | Lead | CRPS | AUC | Trigger (%) | TPR (%) | FPR (%) | Brier Score |
| --- | --- | --- | --- | --- | --- | --- | --- |
| $M_1$ | 1 | 312.8 | 0.73 | 43 | 64 | 32 | 0.206 |
| $M_1$ | 3 | 316.6 | 0.72 | 37 | 65 | 34 | 0.211 |
| $M_1$ | 6 | 330.3 | 0.69 | 31 | 65 | 37 | 0.225 |
| $M_1$ | 12 | 334.4 | 0.65 | 27 | 60 | 40 | 0.244 |
| $M_2$ | 1 | 318.4 | 0.70 | 30 | 62 | 34 | 0.224 |
| $M_2$ | 3 | 322.8 | 0.69 | 27 | 64 | 39 | 0.231 |
| $M_2$ | 6 | 324.9 | 0.68 | 27 | 61 | 37 | 0.236 |
| $M_2$ | 12 | 332.4 | 0.65 | 26 | 57 | 37 | 0.250 |
| $B_1$ | 1 | 336.9 | 0.69 | 43 | 65 | 38 | 0.220 |
| $B_1$ | 3 | 345.4 | 0.67 | 38 | 63 | 41 | 0.227 |
| $B_1$ | 6 | 359.7 | 0.63 | 33 | 59 | 42 | 0.240 |
| $B_1$ | 12 | 362.9 | 0.57 | 29 | 55 | 47 | 0.260 |
| $B_2$ | 1 | 353.4 | 0.62 | 30 | 61 | 45 | 0.242 |
| $B_2$ | 3 | 353.8 | 0.62 | 30 | 61 | 46 | 0.243 |
| $B_2$ | 6 | 354.4 | 0.61 | 31 | 58 | 43 | 0.244 |
| $B_2$ | 12 | 359.9 | 0.58 | 29 | 57 | 47 | 0.256 |

Climate-informed models improved all three metrics relative to their corresponding baseline model at every lead time (Figure 3; Table 2). Comparing M_1_ with B_1_, improvements were consistent across leads for case estimation (7.2-8.4% CRPS) and probabilistic accuracy (6.1-7.1% BS), but outbreak detection improvements widened with lead time, from +0.04 at lead 1 to +0.08 at lead 12. When excluding the yearly random effect (M_2_ with B_2_), improvements narrowed with lead time for case estimation, from 9.9% to 7.6%, and probabilistic accuracy, from 7.2% to 2.4%, yet the outbreak detection gap remained consistent, ranging 0.07-0.08.

### Health region predictive improvements shift from cases at shorter leads to outbreaks at longer leads

To assess spatial variation in skill relative to a baseline, we compared the Continuous Ranked Probability Skill Score (CRPSS), Brier Skill Score (BSS), and difference in AUC (ΔAUC) of the best-fitting model (M_1_) relative to its corresponding baseline model (B_1_) for each health region (Figure 4). The proportion of health regions (n = 435) where climate information improved skill differed by metric and lead time. Case estimation improved skill in 74.7% of health regions at lead 1, declining to 63.9% at lead 12. Similarly, probabilistic accuracy improvements decreased from 73.1% to 65.7% of health regions across lead times. Conversely, outbreak detection improvements increased from 61.4% at lead 1 to a maximum of 71.5% at lead 6, then decreased to 66.9% at lead 12. Predictive skill improvements per health region therefore shifted from case estimation at shorter lead times to outbreak detection at longer lead times.

**Figure 4.**
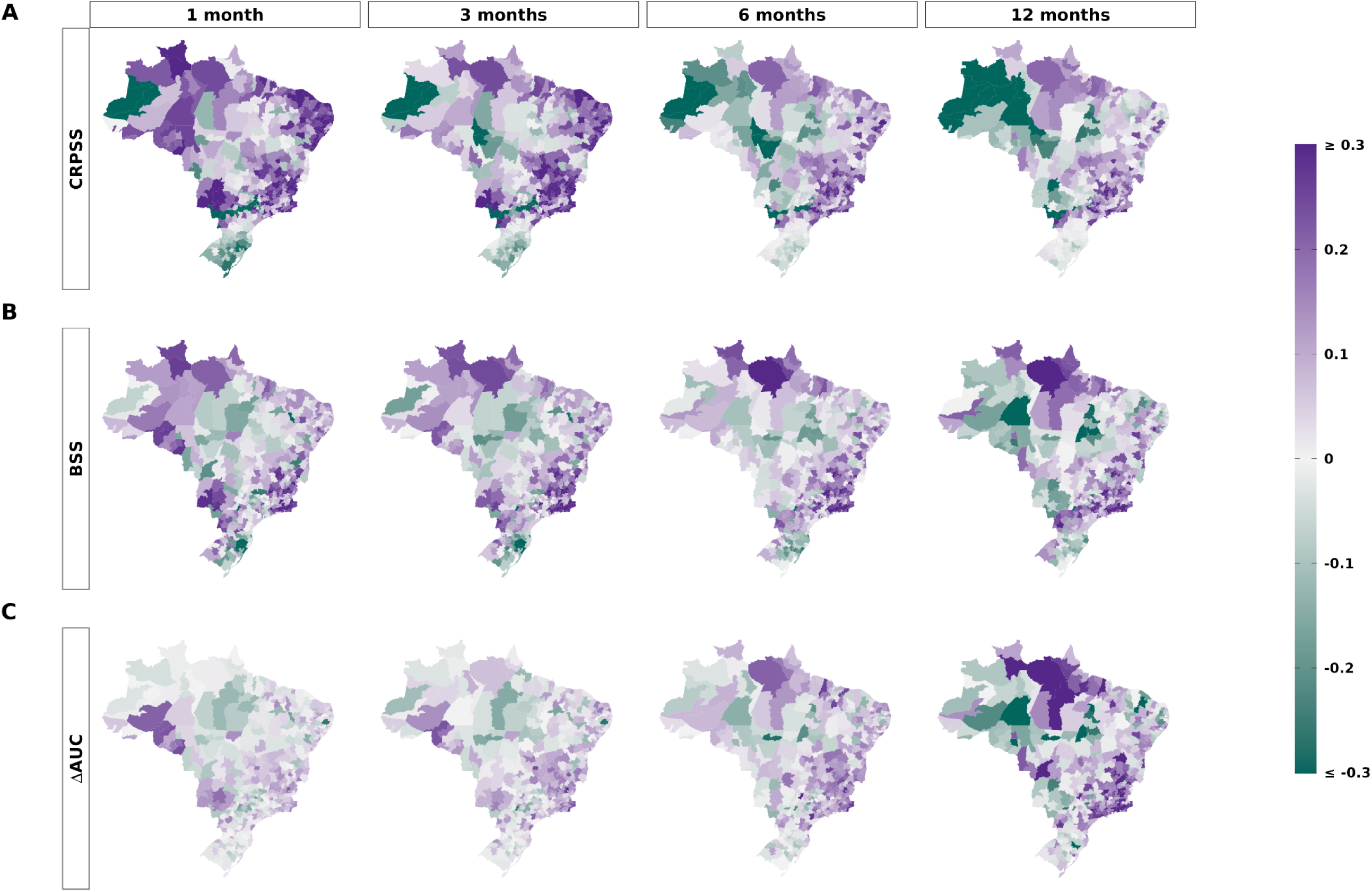
Spatial variation in predictive skill contributed by climate information. Predictive skill of the best-fitting climate-informed model (MJ relative to its baseline (B) for each of the 435 health regions at lead times of 1. 3, 6 and 12 months (columns). (A) Continuous Ranked Probability Skill Score (CRPSS). (B) Brier Skill Score (BSS). and (C) Difference in AUC (ΔAUC). Purple indicates health regions where climate information improved skill, and green indicates where climate information degraded skill. All metrics were capped at ±0.3. Health regions in the state of Espírito Santo are excluded in the maps due to unavailable surveillance data.

Improvements were spatially structured, with the contribution of climate information differing across the five macro-regions. For case estimation, M_1_ outperformed B_1_ in most health regions in the Northeast (67.5-91.5%), Southeast (81.9-85.0%), and Midwest (52.3-77.2%) across all lead times. However, improvements declined rapidly for health regions in the North from lead 1 (82.2%) to lead 12 (37.8%), with skill worsened by climate information in the South at every lead (20.2-37.7%). Probabilistic accuracy showed similar patterns across macro-regions. Outbreak detection skill was less consistent, with improvements for most health regions in the Northeast (59.8-78.6%) peaking at lead 6, Southeast (67.5-87.5%) peaking at lead 12, and Midwest (50.0-65.9%) peaking at lead 1. By contrast, in the North, the AUC improved in only 33.3% of health regions at lead 1, rising to 62.2% at lead 6 and declining to 48.9% at lead 12. The South was split across leads, with 49.3-58.0% showing improvements in outbreak detection with climate information.

### Lagged dengue incidence enhances predictability up to six months ahead

We calculated the CRPS, BS and AUC for two additional models: “DIR” representing the random effects-only model, B_1_, with the lagged dengue incidence rate (DIR), and “Both” representing the climate-informed model, M_1_, with the lagged DIR (Table 3). We evaluated the CRPSS, BSS and ΔAUC relative to B_1_ for three formulations: the best-fitting climate-informed model (M_1_; “Climate”), “DIR”, and “Both” (Figure 5). Here, we included the most recent DIR that would be available per lead time (e.g. lagged 2 months at the 1-month lead). This enabled us to quantify skill improvements from climate information alone, incidence alone, and both climate and incidence combined.

**Figure 5.**
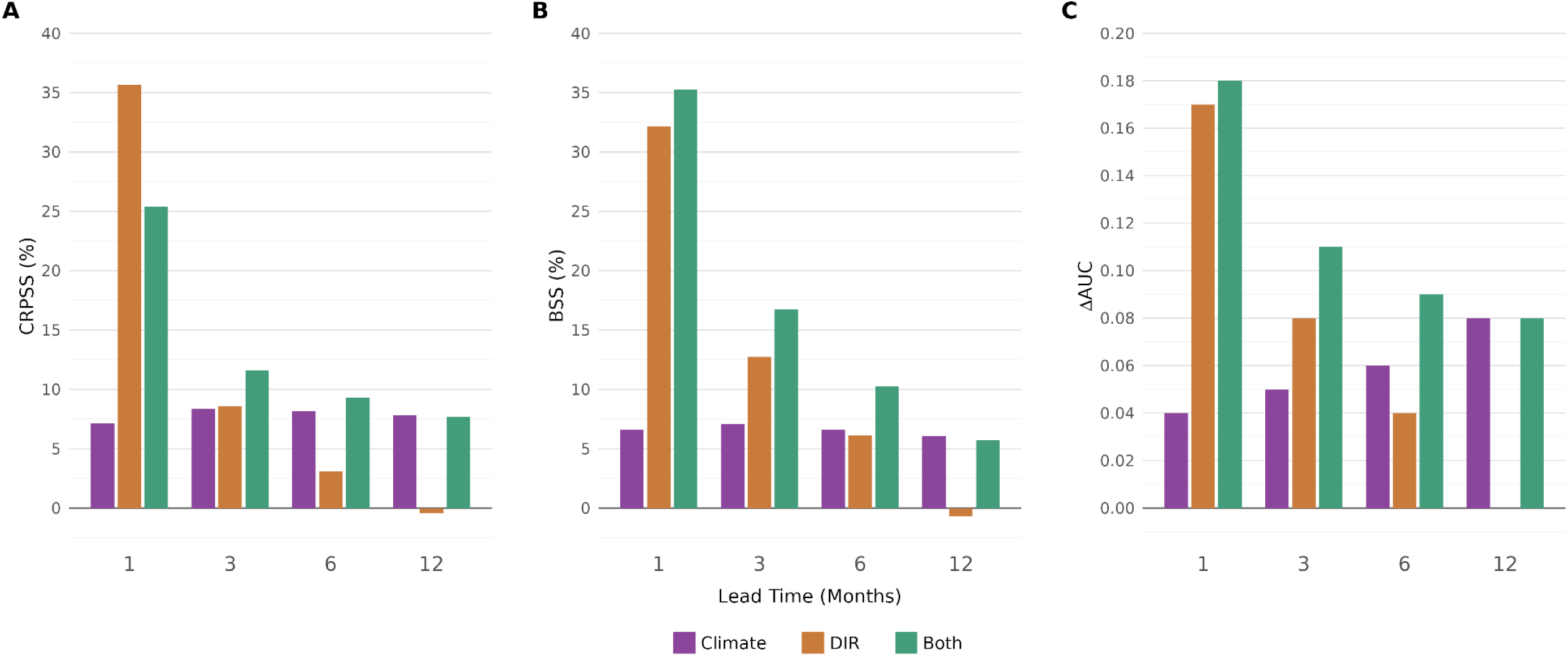
Predictive skill contributed by climate information, lagged dengue incidence, and both combined. Predictive skill relative to the random effects baseline (BJ at lead times of 1. 3, 6 and 12 months, for the best-fitting climate-informed model (M_b_ Climate), a model containing random effects and the lagged dengue incidence rate (DIR), and the climate-informed model with the DIR included (Both). The most recent DIR available at the time of issue was used for each lead time, lagged by the lead time plus one month. (A) Continuous Ranked Probability Skill Score (CRPSS. %). (B) Brier Skill Score (BSS. %). and (C) Difference in AUC (AAUC). Positive values indicate improvement over the baseline.

**Table 3.** Predictive skill for case estimation, probabilistic accuracy and outbreak detection across operational lead times. Out-of-sample predictive skill at lead times of 1, 3, 6 and 12 months, evaluated across all months and health regions using rolling-origin cross-validation with an expanding window from October 2020 to May 2025. Two model formulations were compared: the best-fitting climate-informed model (M_1_) with the lagged dengue incidence rate (DIR) named “Both”, and the corresponding random-effects baseline (B_1_) with the lagged DIR named “DIR”. The lag is relative to the lead time, representing the lead time plus one. The Continuous Ranked Probability Score (CRPS) measures case estimation, the Brier Score measures probabilistic accuracy, the Area Under the Receiver Operating Characteristic Curve (AUC) measures outbreak detection. Lower CRPS and Brier Score, and higher AUC, indicate better skill. Trigger denotes the minimum outbreak probability to raise an alert, derived from the ROC curve as the top-left value. The True Positive Rate (TPR) and False Positive (FPR) are estimated for Trigger. Outbreaks were defined as dengue incidence above the historical 75th percentile for the calendar month and health region.

| Model | Lead | CRPS | AUC | Trigger (%) | TPR (%) | FPR (%) | Brier Score |
| --- | --- | --- | --- | --- | --- | --- | --- |
| $M_1$ + DIR (**Both**) | 1 | 251.3 | 0.87 | 42 | 79 | 21 | 0.143 |
| $M_1$ + DIR (**Both**) | 3 | 305.4 | 0.78 | 39 | 69 | 29 | 0.189 |
| $M_1$ + DIR (**Both**) | 6 | 326.3 | 0.72 | 34 | 63 | 33 | 0.212 |
| $M_1$ + DIR (**Both**) | 12 | 334.4 | 0.65 | 28 | 58 | 38 | 0.245 |
| $B_1$ + DIR (**DIR**) | 1 | 216.7 | 0.86 | 44 | 78 | 22 | 0.149 |
| $B_1$ + DIR (**DIR**) | 3 | 315.8 | 0.75 | 40 | 69 | 32 | 0.199 |
| $B_1$ + DIR (**DIR**) | 6 | 348.6 | 0.67 | 34 | 65 | 40 | 0.226 |
| $B_1$ + DIR (**DIR**) | 12 | 364.4 | 0.57 | 28 | 57 | 49 | 0.262 |

We found that lagged incidence enhances predictive skill up to 6 months ahead, particularly when combined with climate information at lead times of 3 and 6 months. At lead 1, lagged incidence contributed considerably more predictive skill than climate information compared to the baseline, improving case estimation by 35.7% and probabilistic accuracy by 32.2% compared with 7.2% and 6.6% for climate, and increasing the AUC by 0.17 compared to 0.04. This advantage degraded rapidly as lead time increased. Improvements from either climate or DIR alone were comparable between 3-6-month lead times across metrics, whereas DIR provided no gain in predictive skill at the 12-month lead for case estimation (−0.4%), probabilistic accuracy (−0.7%) or outbreak detection (0.00). By contrast, improvements from climate information were stable or increased across leads. Combining both climate and lagged incidence performed best at lead times of 3 and 6 months, where improvements in case estimation (11.6% and 9.3%), probabilistic accuracy (16.7% and 10.2%), and outbreak detection (0.11 and 0.09) exceeded those from either source alone.

### Climate covariates improve monthly predictions of the upcoming dengue season

We simulated long-lead predictions for the upcoming dengue season, forecasting every month over five seasons, starting 2020/21 and ending 2024/25. Each forecast was issued 5 months before the start of the season. We repeated this process for two season definitions, from October to September (Oct-Sep) and from January to December (Jan-Dec), and evaluated predictive skill overall and during the peak from March to May (Figure 6). Evaluations were conducted from January 2021 to May 2025 to compare predictive skill between both definitions (Oct-Sep vs Jan-Dec). We evaluated four models – M_1_, M_2_, B_1_, B_2_ – to further compare whether incorporating climate covariates (M_1_ vs B_1_; M_2_ vs B_2_) or the yearly random effect (M_1_ vs M_2_; B_1_ vs B_2_) improved whole-season or peak-season prediction using the CRPSS, BSS and ΔAUC.

**Figure 6.**
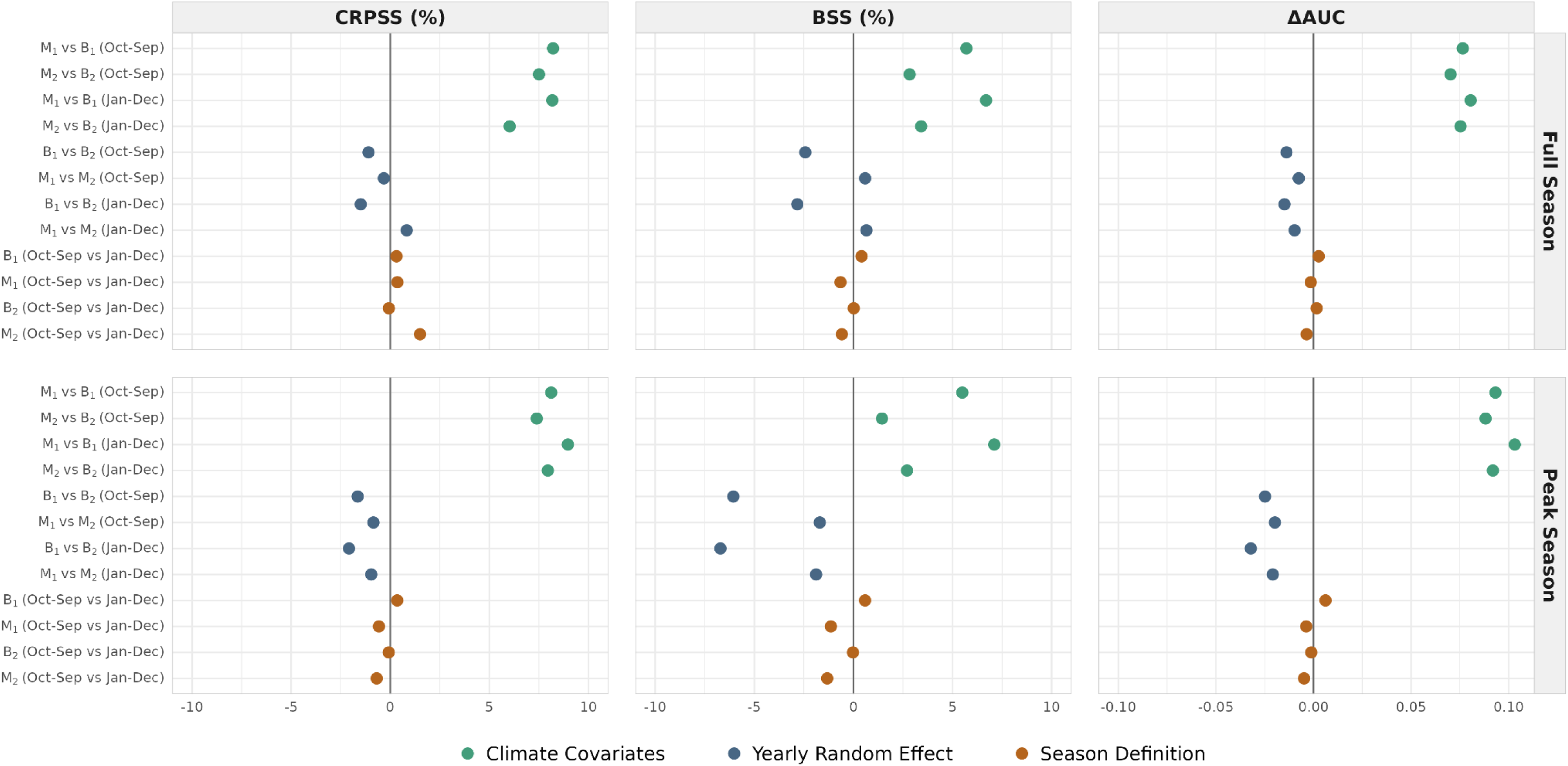
Predictive skill for whole-season dengue forecasting under alternative model and season specifications. Predictive skill for whole-season forecasts issued five months before the start of each season, over five seasons, evaluated from January 2021 to May 2025. Columns show the Continuous Ranked Probability Skill Score (CRPSS, left), Brier Skill Score (BSS, centre) and difference in AUC (ΔAUC, right). Rows show the evaluation over the full season, and peak season from March to May. Each point represents a pairwise contrast in models, with positive values favouring the first model relative to the second model. Three differences are evaluated: Climate Covariates (Mj vs B_b_ M_2_vs B_2_) in green, Yearly Random Effect (Mj vs M_2_, B vs B_2_) in grey, and Season Definition (Oct-Sep, Jan-Dec) in orange. CRPSS and BSS are in percentages, AAUC in AUC units. Observed climate covariates were used at all lead times.

Climate covariates provided the largest and most consistent contribution to predictive skill across all formulations and season definitions (Figure 6). The greatest improvements were exhibited by the best-fitting climate-informed model (M_1_) relative to the baseline (B_1_), particularly during the peak season. M_1_ improved case estimation by 7.4-9.0% for the peak season (6.0-8.2% for full), probabilistic accuracy by 1.4-7.1% for the peak (2.8-6.7% for full), and outbreak detection by 0.09-0.10 for the peak (0.07-0.08 for full).

The yearly random effect predominantly decreased predictive skill, particularly during the peak, although changes were small (Figure 6). Comparing climate-informed models (M_1_ vs M_2_), case estimation and probabilistic accuracy decreased by 0.8-1.9% during the peak season but showed up to 0.8% improvement during the full season, whereas outbreak detection skill consistently degraded by 0.01-0.02. Redefining the dengue season had minimal impact on predictive skill.

### Weekly predictions better capture spatiotemporal distribution than case counts for the season ahead

The monthly modelling framework was extended to the epidemiological week (EW) for the second Infodengue-Mosqlimate Dengue Challenge (IMDC), retaining monthly covariates and adapting the seasonal random effect to the EW level. Three validation seasons – 2022/23, 2023/24 and 2024/25 – were forecast, each issued in EW25 and targeting all weeks from EW41 up to EW40 the following year for 26 federal units, excluding Espírito Santo (Figure 7). To reflect the realistic availability of climate information, climate covariates combined monthly observed data (e.g., reanalysis) until EW25 (May), historical 51-member seasonal forecasts within the 7-month window (June-December), and climatologies thereafter. The yearly random effect was excluded, following improved skill for whole-season prediction at the monthly resolution.

**Figure 7.**
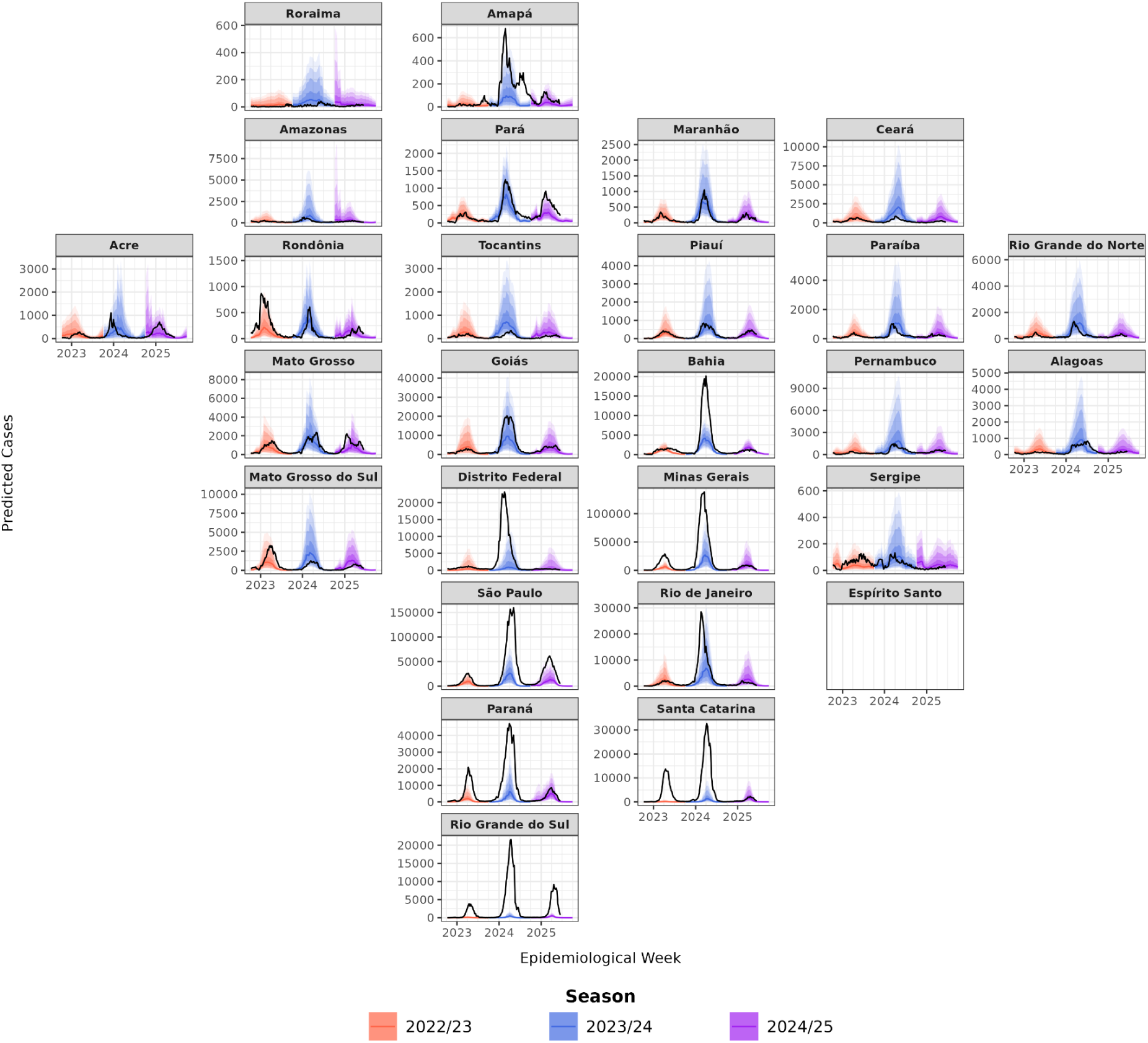
Weekly seasonal forecasts of dengue cases by federal unit. Predicted and observed weekly dengue cases for 26 federal units across three validation seasons (2022/23, 2023/24, 2024/25). We transformed the best-fitting climate-informed model (M_1_) by updating the monthly cyclic random effect to a weekly cyclic random effect on the epidemiological week (EW), and removed the yearly random effect. Each forecast was issued in EW25 and targeted every week from EW41 to EW40 the following year. Black lines show observed weekly cases, with data spanning EW41 2022 to EW25 2025. Solid coloured lines show the median predicted weekly cases, with shaded bands showing the 50%, 80%, 90% and 95% predictive intervals. Climate covariates combined observed ERA5-Land data where the covariate lag exceeded the lead time, SEAS5.1 seasonal forecasts within the 7-month forecast window, and 1991-2020 climatologies at longer leads. Panels are arranged to approximate the geography of Brazil. Note the different y-axis scales per plot. Espírito Santo is shown without observed or predicted cases due to unavailable surveillance records.

The predicted distribution of national cases within each season broadly aligned with observations. The low-transmission period from EW41 to EW52 accounted for 3.6-11.2% of predicted cases compared to 2.3-8.7% of observed cases, whilst the high-transmission period from EW01 to EW25 accounted for 86.1-93.4% compared to 88.6-94.4%. The week of peak national cases was predicted exactly for 2022/23, and within 1 week for both 2023/24 and 2024/25. Spatially, 7 to 8 of the 10 states with the highest observed burden were ranked among the 10 highest predicted in each season. Interannual variation was captured in direction, with the model accurately predicting 2023/24 to have larger case numbers than any season on record.

However, predictions underestimated cases, with 24.8-53.4% of observations captured across seasons. Observed weekly cases were within the 95% predictive interval for 69.5-88.0% of state-weeks, with at least three-quarters of weeks contained within 16 to 22 of the 26 federal units. Agreement was strongest across the Northeast, where observed cases were within the predictive interval for 92.1% of state-weeks, followed by the North (89.4%) and Midwest (85.1%). Predictions were weakest in the Southeast (43.7%) and South (20.3%), where observed cases exceeded the upper bound of the interval in most weeks. Rio Grande do Sul, São Paulo, Santa Catarina, Paraná, and Minas Gerais were the least well-predicted states, despite being among those correctly ranked with the highest burden.

## Discussion

In this study, we quantified the effects of compound climate extremes on dengue risk across 435 health regions of Brazil, and evaluated the predictive skill contributed by climate information at multiple operational lead times from 1 month to over a year in advance. Sequential dry, warm and wet conditions were associated with the highest dengue risk in most climatic zones, with the strength of the temperature anomaly modification differing between tropical and temperate settings. Climate covariates improved case estimation, probabilistic accuracy and outbreak detection at lead times of 1 to 12 months relative to the baseline, and for the whole upcoming season issued 5 months in advance. These improvements were largely concentrated in health regions across the Northeast, Southeast and Midwest of Brazil. Incorporating recent dengue incidence as a covariate improved predictive skill up to 6 months in advance. Extension of the framework to the weekly resolution broadly captured the timing and spatial distribution of dengue cases across federal units, demonstrating adaptability and alignment with operational surveillance and forecasting in Brazil.

Dengue risk increased following a compound sequence of long-lag dry, mid-lag warm and short-lag wet conditions across Brazil, representing the maximum relative risk quantified in 5 of 9 climatic zones comprising 61.6% (268 of 435) health regions. The same compound sequence increased outbreak risk nationally in Barbados (Fletcher et al., 2025). In Brazil, this dry-warm-wet association was predominant in tropical zones (Af, As, Aw), although modification by the temperature anomaly was weak. Interacting long-lag dry and short-lag wet extremes increased dengue risk across all temperature anomalies, indicating that hydrometeorological dynamics may be influential on dengue across tropical settings. Sequential dry-then-wet conditions were similarly associated with the maximum dengue relative risk in seven of eight subregions of Vietnam (Fletcher et al., 2026). Mean temperatures across tropical climates in Brazil remain within the thermal limits for transmission by *Aedes aegypti* mosquitoes (Mordecai et al., 2019), meaning anomalies shift conditions within, instead of outside, the transmission range. By contrast, temperature anomalies strongly modified hydrometeorological dynamics in temperate zones, located in southern Brazil, where dengue risk increased under warmer conditions. Temperature suitability historically constrained transmission across southern Brazil, however, prolonged thermal anomalies under climate change have shifted the geographic barrier southward (Barcellos et al., 2024; Lee et al., 2021). Temperate zones with a dry winter (Cwa, Cwb) demonstrated the same dry-warm-wet sequence, whereas temperate zones without a dry season (Cfa, Cfb) showed maximum relative risk under warmer and persistently dry conditions. This work builds on previous research quantifying risks associated with individual hydrometeorological hazards in Brazil (Lowe et al., 2021) by estimating the effects of concurrent and sequential climate extremes on dengue risk.

Climate covariates improved predictive skill relative to the baseline at every lead time from 1 to 12 months. Improvements in case estimation and probabilistic accuracy remained consistent across leads, whereas improvements in outbreak detection increased with lead time. This indicates that climate information substantially strengthens discrimination between outbreaks and non-outbreak months up to a year in advance, providing ample lead time to inform public health planning such as vector control campaigns, diagnostic and clinical capacity, community mobilisation, and pre-positioning of funding (Alcayna et al., 2025; Díaz et al., 2024). The yearly random effect improved model fit but degraded predictive skill at longer lead times. With limited observations from the season under prediction, the yearly effect reverts toward its prior distribution increasingly as the predictive window expands. This highlights that random effect structures selected on goodness-of-fit do not necessarily improve predictive performance, consistent with findings in Vietnam (Fletcher et al., 2026). The contribution of lagged dengue incidence demonstrated a similar dependence on lead time. Lagged incidence exceeded climate covariates as a source of skill at the 1-month lead. However, this predictive advantage rapidly decreased over time, worsening skill at the 12-month lead. Recent incidence characterises the state of transmission, which embed multiple determinants of disease risk beyond climate alone. Incorporating incidence into the predictive framework depends on reporting delays to the Notifiable Diseases Information System (SINAN). Notification is mandated within seven days of identification, but fewer than half of cases are reported within one week, with an estimated delay of up to 10 weeks (Bastos et al., 2019). In practice, nowcasting of cases via the Infodengue platform could be used as alternative input within the modelling framework to compensate for reporting delays, but these predictions carry additional uncertainty (Bastos et al., 2019; C. Codeco et al., 2018). Combining incidence with climate information exceeded predictability provided by either source alone at lead times of 3 and 6 months, spanning the horizon over which preventative interventions can be realistically planned and carried out. Comparable findings were reported across Colombia, where autoregressive models were skilful at the 1-month lead, and hydroclimatic models at the 3- and 6-month leads (Beal et al., 2025).

Predictive improvements from climate-informed models were spatially heterogeneous across Brazil. Health regions in the Northeast and Southeast demonstrated enhanced skill across all metrics and lead times, with modest gains in the Midwest. These macro-regions have historically experienced the highest and most sustained dengue burden in Brazil with recurrent annual epidemics (Barcellos & Lowe, 2014; Lee et al., 2021). In contrast, climate covariates consistently degraded skill in the South, and skill rapidly declined from lead 1 to 12 in the North. These two macro-regions represent frontiers of recent dengue expansion, where transmission has become established through rapid urbanisation, vector redistribution, population susceptibility, and connectivity with endemic areas (Barcellos et al., 2024; Lee et al., 2021). These non-climatic factors are captured indirectly through spatiotemporal random effects, including the yearly effect, which contributes progressively less at longer lead times. Spatial variation in forecast skill has previously been observed in Brazil, where model performance during the first edition of the IMDC differed between states and seasons, with no single model performing best throughout (Araujo et al., 2026).

Climate information enhanced monthly prediction of the upcoming season issued 5 months ahead, with the greatest improvements during the peak transmission period. Redefining the season to the calendar year had minimal effect on predictive skill, despite shifting the issue date closer to the peak, indicating the stability of climate-driven skill across lead times. However, these evaluations used observed covariates, assuming conditions were perfectly forecasted, representing an upper bound of operational skill (Colón-González, Bastos, et al., 2021). In practice, seasonal forecast uncertainty would propagate through the predictive model, with skill varying by season, location, variable and lead time (White et al., 2017). We incorporated this uncertainty when translating the framework to the weekly resolution used by Brazilian arbovirus surveillance, where cases are notified by epidemiological week and forecasting targets for the Ministry of Health are specified as weekly case curves (Araujo et al., 2026). Depending on lead time and lagged associations, we combined climate observations, historical seasonal forecasts across all 51 ensemble members, and climatologies to reproduce the information available at the time of issue to inform the full predictive distribution. Weekly predictions broadly captured the timing of the peak, spatial distribution of cases, and direction of interannual variation, including the 2023/24 season which recorded the highest number of cases to-date in Brazil (Gurgel-Gonçalves et al., 2024; Haider et al., 2025). However, cases were underestimated in every season, with observed counts exceeding the upper predictive interval in most weeks across the South and Southeast. Relative to the preceding decade, annual incidence in these regions increased 7-fold and 4-fold, respectively, compared with less than a 2-fold increase in the Midwest and stable incidence in the North and Northeast, representing unprecedented surges in infection during the validation period.

These findings indicate that a multi-staged forecasting design could serve operational needs in Brazil. Weekly nowcasts via the Infodengue platform provide timely situational awareness of current dengue transmission that is highly valuable for surveillance (C. Codeco et al., 2018), whereas the IMDC produces multi-model ensemble forecasts of weekly dengue cases over the upcoming season with advanced warning (Araujo et al., 2026). Ensembles can offset the limitations of individual model formulations, and generate more accurate and consistent predictions of dengue risk, particularly across heterogeneous settings (Araujo et al., 2026; Colón-González, Bastos, et al., 2021; Johansson et al., 2019). Furthermore, whole-season predictions aid decision-making with long implementation times, although skill is highly variable across seasons and locations (Araujo et al., 2026). Monthly dengue predictions with 1- to 6-month outlooks could bridge the gap between nowcasting and whole-season forecasting, reducing uncertainty in predicted case counts and outbreak probabilities by re-estimating outcomes with observations or skilful climate forecasts as they become available (Lowe et al., 2018). Progressively updated forecasts would allow preparedness actions to be scaled as targets approach, from resource allocation and procurement at longer leads, to targeted vector control and clinical surge capacity at shorter leads (Alcayna et al., 2025). Our study indicates that recent incidence may be sufficient for predicting dengue risk at the 1-month lead time, whereas combining climate information with incidence improved prediction at the 3- and 6-month leads. Existing nowcast models could be adapted to estimate recent monthly incidence within this framework to compensate for reporting delays. Further evaluation will be required to ensure nowcasted incidence and associated uncertainty retains the predictive gains exhibited by observed incidence.

Several limitations should be considered when interpreting these findings. Dengue cases were defined as probable, which may include misclassified cases of chikungunya and Zika virus (Oliveira et al., 2020). Due to high rates of asymptomatic infection, dengue data represents only symptomatic or notified infections, which are affected by local outbreaks, healthcare access, and surveillance or testing capacity (Bhatt et al., 2013; Coelho et al., 2016). Other important determinants of dengue risk, such as population immunity, vaccination campaigns, serotype switches, and connectivity (Lee et al., 2021; Lorenz et al., 2026; Nogueira et al., 2024; Ranzani et al., 2026; Rodriguez-Barraquer et al., 2011), were represented indirectly through random effects which limit the interpretation of effects. Health region level Köppen-Geiger zones were assigned from the dominant classification across municipalities and assumed stationary over the study period, masking finer-scale heterogeneity and boundary shifts due to climate change (Beck et al., 2023).

Nonetheless, our framework evaluates the effects of concurrent and sequential climate extremes on dengue risk, building on previous research in Brazil (Lowe et al., 2021). We extend the temporal, three-way compound extremes framework in Barbados (Fletcher et al., 2025), and spatiotemporal two-way compound extremes framework in Vietnam (Fletcher et al., 2026), capturing the effects of temperature anomalies, and long-lag and short-lag drought indices on dengue risk across distinct Köppen-Geiger zones in Brazil. This framework could be adapted to assess the effects of concurrent and sequential climate hazards or drivers on disease risk across diverse geographical settings. Furthermore, these findings could support early warning initiatives to enhance public health preparedness and response in Brazil.

## Materials and Methods

### Study area

Brazil is the largest country in South America, both by land area, 8.5 million km^2^ (IBGE, 2025), and population size, 214.2 million people (IBGE, 2026). The country is administratively divided into five macro-regions (North, Northeast, Midwest, Southeast, and South), 27 federal units, and 5570 municipalities. To facilitate the management and integration of health services, municipalities are grouped into 450 health regions (Regiões de Saúde), which serve as the primary spatial unit of analysis for this study. Due to reporting issues, epidemiological data for the state of Espírito Santo were not available, reducing the number of health regions in our study to 435 (Figure 1).

### Dengue data

Dengue surveillance data were obtained from the Notifiable Diseases Information System (SINAN), a national, freely accessible database managed by the Brazilian Ministry of Health. Monthly health region level dengue case counts were obtained using the microdatasus R package (version 2.4.3) (Saldanha, 2026) and processed with the data4health R package (version 0.1.1) (Lührsen et al., 2026), whereas weekly municipality level data were obtained via the second edition of the Infodengue-Mosqlimate Dengue Challenge (IMDC) (Mosqlimate, 2026). Notification of dengue cases is mandatory in Brazil, requiring healthcare professionals to report all suspected cases. A suspected case is defined as acute fever of two to seven days accompanied by at least two of the following: nausea, vomiting, rash, myalgia, arthralgia, headache, retro-orbital pain, petechiae, a positive tourniquet test, or leukopenia (Ministério da Saúde, 2024a). Notified cases are subsequently classified as probable or discarded following investigation, with confirmation based either on laboratory testing (NS1 antigen detection, positive viral isolation, RT-PCR, or IgM serology via ELISA) or on clinical-epidemiological criteria. We used probable cases, which include all notified cases except discarded cases (Ministério da Saúde, 2024b). We aggregated dengue cases per health region at both monthly and weekly resolutions from January 2010 to May 2025. We also obtained annual population estimates per municipality from DataSUS (DataSUS, 2025) and aggregated to health regions to calculate dengue incidence rates (DIR).

### Climate covariate data

We derived a range of monthly temperature and precipitation indices across rolling 1-, 3-, 6- and 12-month windows by spatially averaging data from the high-resolution (0.1°) ERA5-Land global reanalysis dataset (Muñoz-Sabater et al., 2021). Temperature indices included mean, minimum and maximum temperatures (°C), as well as the standardised mean temperature anomaly, defined as standard deviations from the health region-specific monthly climatology using the reference of 1981-2010. Precipitation indices included total precipitation (mm), the Standardised Precipitation Index (SPI), and the Standardised Precipitation Evapotranspiration Index (SPEI). SPI characterises meteorological drought by calculating rainfall anomalies against historical precipitation data using a gamma-fitted distribution. SPEI is also a drought index that accounts for rainfall and potential evapotranspiration with historical precipitation and temperature data using a log-logistic distribution. For both indices, negative values indicate drier-than-normal conditions and positive values indicate wetter-than-normal conditions, respectively, relative to the reference of 1950-2010. We obtained data for the monthly averaged sea surface temperature (SST) in the Niño 3.4 region (5N-5S, 170W-120W) of the Pacific Ocean from the National Oceanic and Atmospheric Administration ERSSTv5 dataset, and calculated the anomaly using the reference of 1981-2010. Rolling means were calculated over 3, 6 and 12 months as trailing averages to preserve temporal precedence (e.g. the 3-month mean for March includes observations from January, February and March). All temperature, precipitation and oceanic indices were lagged 0 to 6 months.

### Climatic zone and biome data

To investigate heterogeneity in the climatic drivers of disease, we defined various spatial and temporal classification variables (Figure 1). We obtained municipality-level Köppen climate classifications (Alvares et al., 2013) and biome classifications (Silva et al., 2022) from the second edition of the IMDC (Mosqlimate, 2026). We assigned each health region the dominant climate type across the respective municipalities, representing 9 classifications including 3 tropical climates (Af: tropical rainforest, Am: tropical monsoon, As: tropical savanna with a dry summer, Aw: tropical savannah with a dry winter), 1 semi-arid climate (BSh: hot semi-arid), and 3 temperate climates (Cfa: humid subtropical, Cfb: temperate oceanic, Cwa: dry-winter humid subtropical, Cwb: subtropical highland). Similarly, we assigned each health region the dominant biome type across the respective municipalities, representing 6 classifications including the Amazon (Amazônia), Savanna (Cerrado), Desert (Caatinga), Atlantic Forest (Mata Atlântica), Grassland (Pampa), and Flooded Savanna (Pantanal) biomes.

### Model formulation

In this study, we developed climate-informed disease risk models from January 2010 to May 2025, defining a dengue season from October to September. Models were developed at the health region level, *s* (1 to 435), and monthly timescale, *t* (1 to 185). We specified Bayesian hierarchical models using dengue case counts, *y_s,t_*, as our response variable. Case counts were assumed to follow a negative binomial distribution to account for potential overdispersion such that:

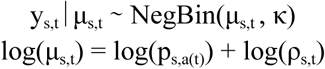

with distribution mean, *μ_s,t_*, and overdispersion parameter, *κ*. The distribution mean represents the population, *p_s,a(t)_*, in a given health region, *s*, and year, *a(t)*, multiplied by the dengue incidence rate, *ρ_s,t_*. The logarithm of population was therefore used as a model offset.

Firstly, we developed a baseline model that included spatiotemporal random effects and a global intercept, *α*. To account for seasonal variation in dengue incidence, we defined a temporal random effect, *δ_f(s),m(t)_*, for each calendar time point, *m(t)*, from 1 to 12 months, and evaluated the seasonality nationally or replicated by federal unit, macro-region, biome, Köppen climate zone, represented as *f(s)*. The seasonal effect was specified as a cyclic second-order (RW2) random walk model. In years with 53 epidemiological weeks, week 53 was assigned to week 52. To account for interannual variation in dengue incidence, we defined a temporal random effect, *γ_g(s),a(t)_*, for each year, *a(t)*, from 1 to 16, and evaluated nationally or replicated by federal unit, macro-region, biome or climate zone, represented functionally as *g(s)*. The yearly effect was specified as an independently and identically distributed (IID) model. To account for spatial variation in dengue incidence, we defined a spatial random effect with structured, *u_s_*, and unstructured, *v_s_*, components by health region, *s*. The spatial effect was specified as a modified Besag-York Mollié (BYM2) model. The baseline model was therefore formulated such that:

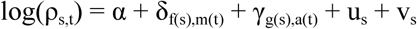

### Climate-informed model selection

Building upon the baseline model, we incorporated covariates through a stepwise selection process to determine key drivers of dengue incidence. We first conducted a univariable analysis for all lagged climate covariates, estimating linear and nonlinear effects, alongside random slopes to stratify effects by macro-region, Köppen-Geiger climate zone, and biome classification. For nonlinear effects, the range of each covariate was divided into 10 equal-width intervals, with an effect estimated for each interval using an RW2 prior to smooth the relationship between neighbouring intervals. We assessed goodness-of-fit using five metrics: the Deviance Information Criterion (DIC), Watanabe-Akaike Information Criterion (WAIC), Log Mean Score (LMS), Mean Absolute Error (MAE), and Root Mean Squared Error (RMSE). Covariates with maximised fit, decreased random effects, and fixed-effect estimates whose 95% credible intervals excluded zero, were combined into additive multivariable models, testing combinations with two to four covariates.

We expanded the best-performing multivariable models to capture compound climate extremes. For this, we formulated a three-way interaction between temperature indicators (*X_T_*), and long-lag (*X_L_*) and short-lag (*X_s_*) precipitation indicators, with individual effects (*β_T_*, *β_L_*, *β_S_*), pairwise interactive effects (*β_T,L_*, *β_T,S_*, *β_L,S_*), and a trivariable interactive effect (*β_T,L,S_*). We evaluated the interaction linearly and with random slope stratification of effects by macro-region, Köppen-Geiger climatic zone, and biome, represented by *r(s)*. Additive terms were represented by *X_k_* with corresponding *β_k_* coefficients. The interaction model was formulated such that:

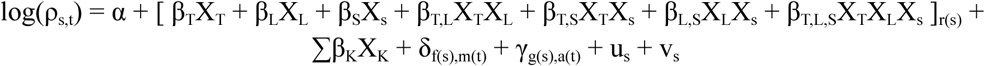

Model parameters were estimated in R version 4.6.1 using the GHRmodel package version 0.1.1 (Milà et al., 2025) with the Integrated Nested Laplace Approximation (INLA) version 23.04.24 (Rue et al., 2009).

### Predictive skill evaluation

We assessed the predictive skill of the best-fitting monthly model using rolling-origin cross-validation with an expanding window from October 2020 to May 2025 at lead times of 1, 3, 6 and 12 months. For each prediction month, models were fitted exclusively on historical data, withholding data points within the set lead time and an additional month to account for a reporting delay. We expanded the training window by one month for each successive prediction. Predictive skill of the best-fitting model was compared with the baseline model, and model formulations with small changes, such as removing the yearly random effect or adding the DIR, lagged *l* + 1 for lead time, *l*, as a covariate.

Predictive skill was evaluated using the Continuous Ranked Probability Score (CRPS) for case estimation, the Brier Score (BS) for the accuracy of probabilistic predictions, and the Area Under the Receiver Operating Characteristic Curve (AUC) for outbreak detection. An outbreak was defined as a month in which observed dengue incidence exceeded the 75th percentile of the historical distribution for the same calendar month and health region, calculated using the training data. For comparing models, we used the Continuous Ranked Probability Skill Score (CRPSS), Brier Skill Score (BSS), and difference in AUC (ΔAUC).

### Operational prediction of the dengue season

Operational public health planning in Brazil requires anticipating the whole dengue season several months before it commences (Araujo et al., 2026). To evaluate it, we adapted the workflow to carry out whole-season prediction using the same monthly dataset, fitting the model to historical data and predicting all months of the upcoming season simultaneously. Two season definitions were evaluated: October to September (dengue epidemiological season), and January to December (calendar year). Forecasts were issued five months before the start of each season, with multi-lead targets from 5 to 16 months ahead. Five seasons were predicted under each definition, beginning October 2020 or January 2021, with the final season truncated at May 2025. For comparability between definitions, predictive skill was evaluated over the common window from January 2021 to May 2025. We assessed predictive skill for the whole season as well as the peak season (i.e. March to May). We compared models with and without covariates, and with and without the yearly random effects, using the CRPSS, BSS and ΔAUC. Therefore, our evaluation spanned four dimensions: (i) season definition, (ii) climate covariates, (iii) yearly random effect, and (iv) whole vs peak season. Climate covariates used observations at all lead times, treating climate as perfectly forecast, to reflect skill attributable to the model without seasonal forecast uncertainty.

We extended the predictive modelling framework to the epidemiological week (EW) resolution, at which dengue surveillance and operational forecasting are routinely conducted in Brazil (Araujo et al., 2026; C. Codeco et al., 2018). The model structure was retained, with monthly covariates assigned to that of fourth day of the epidemiological week (i.e. Wednesday), and the seasonal random effect redefined from the calendar month, m(t) to the epidemiological week, w(t), from 1 to 52. In years with 53 epidemiological weeks, week 53 was assigned to week 52. Each forecast targeted the season from EW41 to EW40 the following year, issued in EW25. We aggregated health region predictions to the federal unit level, and quantified case numbers per state with a range of percentiles (2.5th, 5th, 10th, 25th, 50th, 75th, 90th, 95th, 97.5th). This design follows the Infodengue-Mosqlimate Dengue Challenge (IMDC), in which models are validated against past seasons before forecasting the upcoming season to contribute to an ensemble model used to aid the Brazilian Ministry of Health with public health planning (Mosqlimate, 2026). The weekly dengue data were obtained from the second edition of the IMDC. To make operationally relevant predictions, climate covariates for the weekly forecasts combined three sources according to data availability at the time of issue. Observed ERA5-Land data were used up to EW25 (May). Beyond the observed period, covariates were obtained from the European Centre for Medium-range Weather Forecasting (ECMWF) SEAS5.1 seasonal forecast system (Johnson et al., 2019), using the 51-member forecast initialised in June of each season, for up to 7 months lead (i.e. June to December). Forecasts were calibrated against reanalysis over a 1993-2016 hindcast period, applying member-by-member variance inflation (EVMOS) to temperature and SST variables, and empirical quantile mapping to precipitation using the CSTools package (Perez-Zanon et al., 2026). Covariates were calculated separately for each of the 51 ensemble members, and predictions were generated for each member, with posterior predictive samples pooled across members to form the predictive distribution. Beyond the seasonal forecast window, covariates were set to climatologies over the 1991-2020 reference period.

## Data availability

Dengue surveillance data were obtained from the Notifiable Diseases Information System (SINAN) database, a national freely accessible database managed by the Brazilian Ministry of Health. Monthly health region level dengue case counts were obtained using the microdatasus R package (version 2.4.3) (Saldanha, 2026) and processed with the data4health R package (version 0.1.1) (Lührsen et al., 2026). Weekly dengue case count data per municipality, along with annual population, Köppen-Geiger climate zones and biome classifications per municipality, were obtained from the second edition of the Infodengue-Mosqlimate Dengue Challenge (IMDC; https://sprint.mosqlimate.org/). Temperature and precipitation data were obtained from the ERA5-Land reanalysis dataset (https://cds.climate.copernicus.eu/datasets/reanalysis-era5-land), and oceanic indices from the NOAA Physical Sciences Laboratory (https://psl.noaa.gov/data).

## Acknowledgements

We thank Dr Raquel Lana from the Barcelona Supercomputing Center (BSC) for valuable discussions throughout the study. We acknowledge support from the Wellcome Trust under three separate projects: IDExtremes (grant number 226069/Z/22/Z), Mosqlimate (grant number 226088/Z/22/Z), and HARMONIZE (grant number 224694/Z/21/Z). D.L. was supported by the Barcelona Supercomputing Center AI4Science Fellowship programme funded by the Recovery and Resilience Mechanism-Next Generation as part of the Spanish Ministry’s Recovery, Transformation and Resilience Plan

